# A multi-trait approach identified seven novel genes for back pain-related phenotypes

**DOI:** 10.1101/2024.06.16.24309012

**Authors:** Nadezhda M. Belonogova, Elizaveta E. Elgaeva, Irina V. Zorkoltseva, Anatoliy V. Kirichenko, Gulnara R. Svishcheva, Maxim B. Freidin, Frances M. K. Williams, Pradeep Suri, Tatiana I. Axenovich, Yakov A. Tsepilov

## Abstract

Back pain (BP) is a major contributor to disability worldwide. We conducted a cross-sectional study analyzing three BP-related phenotypes: chronic BP (CBP), dorsalgia and intervertebral disc disorders (IDD), with heritability estimated at 40-60%. Less than half of the heritability is explained by common genetic variants identified by GWAS. More powerful methods of statistical analysis may offer additional insight. Using imputed genotypes from the UK Biobank we performed a multi-trait gene-based association analysis of three BP-related phenotypes: CBP, dorsalgia, and IDD. We identified and replicated 16 genes associated with BP-related traits. Seven of the detected genes, namely, *MIPOL1, PTPRC, RHOA, MAML3, JADE2, MLLT10*, and *RERG*, were previously unreported. Several new genes have been previously detected as associated with traits genetically correlated with BP or as included in pathways associated with BP. Our results verify the role of these genes in BP-related traits and provide new insights into the genetics of back pain.

## 1. Introduction

Back pain (BP) is an important contributor to global disability [1,2]. Chronic back pain (CBP), defined as self-reported back pain lasting more than three months, is the most debilitating form of back pain. It is prevalent in approximately 10%, depending on the population and the definition used [2]. CBP is a complex heritable trait with heritability ranging from 40 % to 68 % [3]. Genes associated with CBP have been identified in several genome-wide association studies (GWAS) using data from national biobanks [4-8]. Recent advances in BP genetics research have been made by meta-analysis of other BP-related phenotypes such as dorsalgia and intervertebral disc disease (IDD) [6]. A total of 41 variants at 33 loci were identified by the authors. However, the liability scale heritability of BP estimated from the SNPs is only 13%, which is significantly lower than the 40% heritability of BP expected from classical twin studies [9]. Several explanations have been proposed to explain this lack of heritability. One of them is the need for more powerful methods of association analysis.

In this study, we utilized a novel multi-trait approach to merge three BP-related phenotypes: CBP, dorsalgia, and IDD, which were previously used in BP studies. Dorsalgia is a phenotype based on electronic health records (EHR) and is defined by clinical diagnostic codes for back and neck pain, reflecting mainly the former. IDD is another phenotype related to BP and defined by EHR-based codes for intervertebral disc degeneration. These codes are typically used when BP is present, but not necessarily [4,10]. There are high genetic correlations between BP-related phenotypes, estimated to be in the range of 87% to 92% [4,11], allowing the use of methods that maximize multi-trait heritability. The aim of this approach is to reduce the genetic heterogeneity of the traits and to identify a common genetic background for a number of traits that are genetically correlated [12].

We also used gene-based association analysis, which is a robust method because it estimates the combined effect of a set of variants within a gene, rather than analyzing each variant individually [13]. This method enables the identification of genes whose protein-coding mutations alter the structure of the corresponding proteins or whose intragenic mutations affect gene expression. Recently, we applied multi-trait and gene-based approaches to analyze the association between BP and rare exome-sequenced variants and identified a new *FSCN3* gene [4,11].

In this study, our aim was to continue the analysis of the genes responsible for differences in the risk of BP between individuals using multi-trait gene-based association analysis. We did this by testing the effect of common variants typically tested by GWAS, using imputed genotypes from the UK Biobank database.

## 2. Materials and Methods

The study design and the main results are presented in Figure 1.

**Figure 1.**
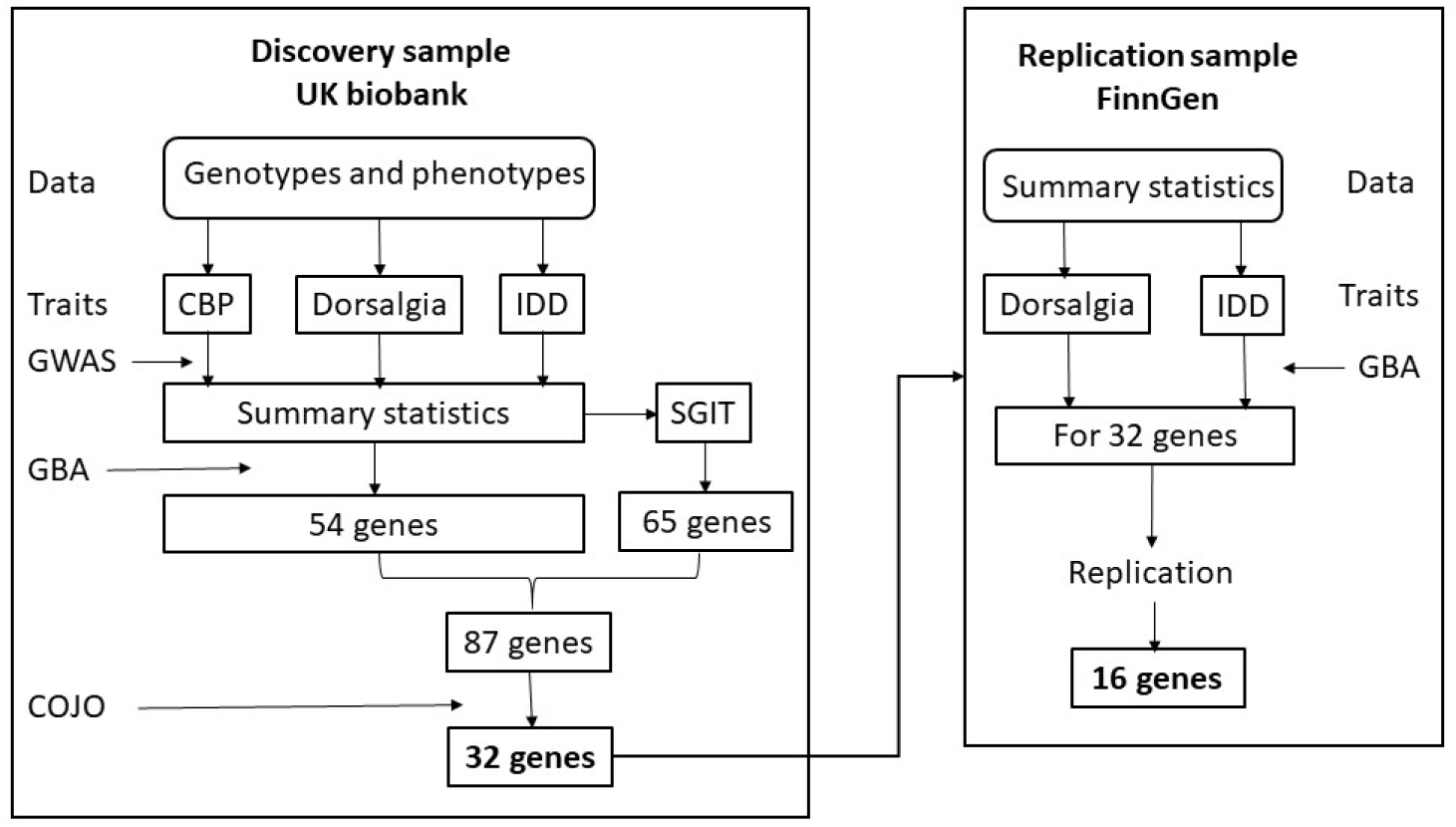
Design of the study and the main results obtained in different steps.

The first step of the analysis was performed using a discovery sample from the UK Biobank. Initially, the GWAS summary statistics for the three BP-related phenotypes were obtained. They were used for the calculation of the multi-trait (SGIT) summary statistics. Then we performed a gene-based association analysis (GBA) of the individual and multi-trait phenotypes. For all identified genes, conditional analysis (COJO) was applied. The second step of the analysis used a replication sample from the FinnGen database. We performed a replication study by applying the GBA to 32 genes identified in the first step and using GWAS summary statistics for two BP-related traits available on the FinnGen sample.

### 2.1 Data

We performed our cross-sectional study using the UK Biobank 500k dataset. The study was done using individuals of white European ancestry. Data field 21000 was used to define self-reported ethnic background.

#### 2.1.1 Phenotypes

The study examined three BP-related conditions: CBP, dorsalgia, and IDD. For CBP, cases and controls were defined on the basis of responses to a questionnaire (data field 3571 “Back pain for 3 months or more”). The details have been described in our previous paper [11] and are presented in Supplementary Methods.

EHR-based phenotypes of dorsalgia and IDD were defined by the level 2 ICD-10 codes M54 and M51, respectively.

For a comprehensive list of ICD-10 codes in the UK Biobank, please refer to https://biobank.ndph.ox.ac.uk/ukb/field.cgi?id=41202.

Numbers of individuals used in the study were 447 466 for CBP and 459,161 for IDD and dorsalgia.

The prevalence of three BP-related phenotypes was 17.97% for CBP, 3.48% for dorsalgia, and 1.76% for IDD. Many individuals have two or three of these BP-related phenotypes simultaneously (Supplementary Figure S1). In particular, 68% of individuals with IDD also have CBP and/or dorsalgia.

#### 2.1.2 Genotypes

We used genotyping and imputation data from the UK Biobank March 2018 data release. Genotypes were obtained using the Affymetrix UK BiLEVE and Affymetrix UK Biobank Axiom arrays and imputed with the IMPUTE4 program (https://jmarchini.org/impute-4/) [14] using the Haplotype Reference Consortium (HRC) [15] and merged UK10K and 1000 Genomes phase 3 reference panels. Genotypes of 19 397 568 SNPs, INFO > 0.8 and MAF > 5×10^-5^ were analyzed.

### 2.2 Gene-based association analyses

For gene-based association analysis, summary statistics (z-scores and effect sizes) for each variant, and the matrices of correlations between genotypes of all variants within a gene were calculated.

#### 2.2.1 Summary statistics

The GWAS summary statistics for each BP-related trait were calculated using the fastGWA-GLMM tool, version 1.94.0 beta [16]. To account for random effects mediated by relatedness, we used a sparse genomic relationship matrix. This matrix was calculated by fastGWA-GLMM (--make-bK-sparse option with default parameters) using the common genetic variants of 487 000 individuals. The analysis was performed using the --fastGWA-mlm-binary option and filters keeping variants with MAF ≥ 5×10^-5^ and variant messiness rate *≤* 0.02. We included sex (data field 22001), age (data field 34), genotyping batch (data field 22828), and the first ten genetic principal components provided by the UK Biobank as covariates in the association analysis to correct for fixed effects. The summary statistics were calculated on the GRCh38/ hg38 genomic build because the gene boundaries and variant annotations are more correct for this build than for previous builds. We used the command line tool liftOver (https://genome-store.ucsc.edu/) and UCSC chain file hg19ToHg38.over.chain.gz (http://hgdownload.soe.ucsc.edu/goldenPath/hg19/liftOver/) to convert genome coordinates from GRCh37/hg19 assembly to GRCh38/hg38 assembly.

#### 2.2.2 Matrices of genotype correlations

For every pair of variants within a gene, we calculated the correlation between their genotypes using 315 599 unrelated white UK Biobank participants. PLINK v2.00a3.7LM (https://www.cog-genomics.org/plink/2.0/) with options –maf 5E-05 --geno 0.02 was used.

#### 2.2.3 Variant annotations

Genetic variants were annotated using the Ensembl Variant Effect Predictor (VEP) [17].

We analyzed three variant annotations: protein coding (exons), protein non-coding (introns, 5′UTR, and 3′UTR), and nonsynonymous SNPs. The latter included transcript ablation, frameshift, stop gained, stop lost, start lost, transcript amplification, inframe insertion, inframe deletion, missense, and protein-altering variants. The numbers of the variants in each variant annotation are presented in Supplementary Table S1.

#### 2.2.4 Methods of gene-based analysis

Three methods based on the summary statistics were applied: SKAT-O [18], PCA [19], and ACAT-V [20]. These methods were implemented in the sumFREGAT R-package [21]. The results of these methods were combined by the aggregated Cauchy omnibus test, ACAT-O [20]. We analyzed protein-coding genes with at least two variations that had summary statistics. The significance level was defined as 2.5×10^-6^.

### 2.3 Multi-trait analysis

The SHAHER framework aims to create a multi-trait phenotype from a set of genetically correlated traits and identify the genetic variants that control this trait [12]. The method assumes that the genetic background of each of the genetically correlated traits can be broken down into two components: one common to all traits (the shared genetic impact or SGI), and the other specific to each trait. A new multi-trait phenotype, SGIT (the shared genetic impact trait), was created by linearly combining the original traits. The coefficients in this combination were defined by maximizing the heritability of SGIT. A GWAS of SGIT was then performed using an algorithm based on the summary statistics calculated for the original traits. The gene-based analysis was performed in the usual way. The SHAHER method was previously described in our studies [11,12] and is explained in detail in the Supplementary Methods.

### 2.4 Conditional analysis

The GCTA-COJO tool was utilized to conduct conditional analysis [22,23]. The variants within 5 Mb of the gene borders were subjected to the COJO selection process. The lowest p-value found in the gene region under examination was designated as the threshold p-value for index variants (--cojo-p). If independent variants chosen at this stage were found to be located outside of the examined gene region, they were considered conditional. Gene-based analysis then used conditional summary statistics (p-values, betas) computed for the variants within the examined gene region.

### 2.5 Replication

GWAS summary statistics for phenotypes of ‘dorsalgia’ and ‘other intervertebral disc disorders’ of the FinnGen release v.9 [24] were used to obtain gene-based p-values for statistically significant genes. Unfortunately, CBP summary statistics were unavailable because they were not included in the FinnGen release v.9. Summary statistics were filtered by INFO > 0.8 and MAF > 5×10^-5^. A replication threshold p-value was calculated as 0.05/(number of genes selected for replication). A gene was considered replicated if the p-value for either ‘dorsalgia’ or ‘other intervertebral disc disorders’ phenotype in the FinnGen sample was lower than the replication threshold.

## 3. Results

### 3.1 Gene-based association analysis of original traits

We observed 86 genome-wide significant gene-based signals across all trait-annotation combinations, with 54 unique genes detected in total: 51 for CBP, three for IDD, and two for dorsalgia (two genes associated with both CBP and IDD, see Supplementary Table S2).

Full results of the gene-based analysis are available in the ZENODO database (https://doi.org/10.5281/zenodo.8118630). QQ-plots for gene-based analyses are shown in Supplementary Figure S2. Manhattan and QQ plots for single-point association analyses are shown in Supplementary Figure S3.

### 3.2 Multi-trait analysis

Using the phenotypic and genetic correlations between the original traits and their SNP-based heritability (Supplementary Figure S4, first three columns and rows), we estimated the coefficients of optimal linear combination of the original traits to build SGIT as 0.69, 0.44, and 0.31 for CBP, dorsalgia, and IDD, respectively. Then, we calculated the phenotypic and genetic correlations between the original traits and SGIT and estimated the SNP-based heritability of SGIT (Supplementary Figure S4, fourth column and row). The phenotypic correlations between SGIT and the original traits are stronger than those between the original traits. All genetic correlations between SGIT and the original traits are no less than 0.93. The SNP-based heritability of SGIT is higher than that of the individual traits.

### 3.3 Gene-based association analysis of SGIT

We observed 105 signals in 65 genes (Supplementary Table S2).

### 3.4 Conditional analysis

We tested a total of 191 signals in 87 genes identified using the original traits and SGIT. After conditional analysis, 15 of 80 signals obtained for CBP retained significance. For IDD, one of four signals passed conditional analysis. For SGIT, 25 of 105 signals retained significance after conditional analysis. For dorsalgia, neither of the two genome-wide significant signals passed COJO (Supplementary Table S2). In total, 32 genes were selected for replication.

### 3.5 Replication

We tested the replication of 32 genes left after the conditional analysis using FinnGen summary statistics obtained for IDD and dorsalgia. The threshold p-value level for replication was calculated as 0.05/32 = 0.00156. A total of 16 genes were replicated (Table 1). All genes were replicated using non-coding variant annotation. The *CHST3* gene identified for IDD passed the replication threshold. Among 15 genes significantly associated with CBP after conditional analysis, two genes, *C8orf34* and *RHOA*, were replicated. Among 24 genes identified for SGIT, 13 genes were replicated. Six of them, *DCC, ILRUN, MAML3, MIPOL1, SOX5*, and *SPOCK2*, were also replicated for CBP (Table 1, Figure 2).

**Table 1.**
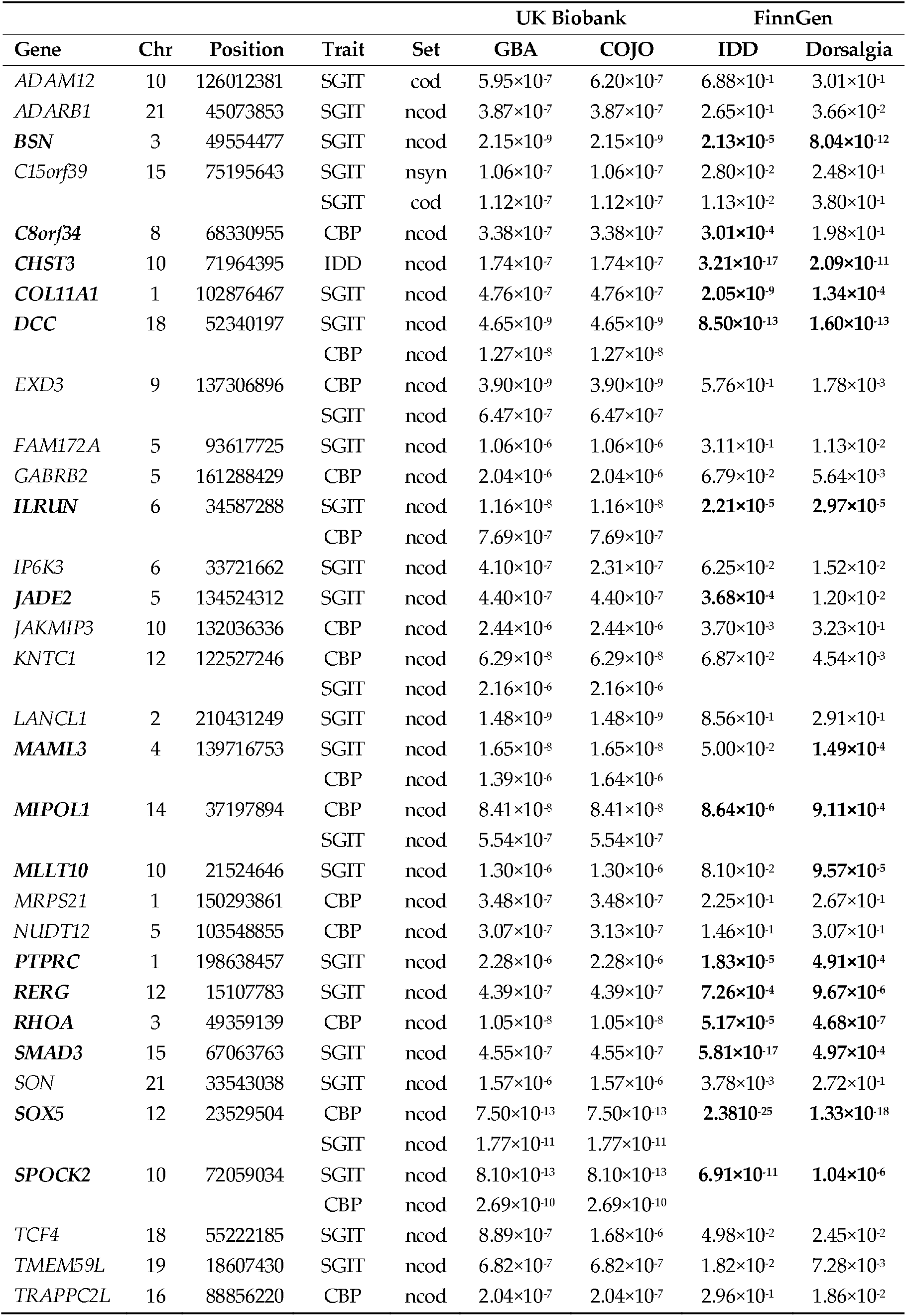
Replication results on FinnGen data. Genes that passed the replication threshold are shown in bold.

| Gene | Chr | Position | Trait | Set | UK Biobank |  | FinnGen |  |
| --- | --- | --- | --- | --- | --- | --- | --- | --- |
|  |  |  |  |  | GBA | COJO | IDD | Dorsalgia |
| *ADAM12* | 10 | 126012381 | SGIT | cod | $5.95 \times 10^{-7}$ | $6.20 \times 10^{-7}$ | $6.88 \times 10^{-1}$ | $3.01 \times 10^{-1}$ |
| *ADARB1* | 21 | 45073853 | SGIT | ncod | $3.87 \times 10^{-7}$ | $3.87 \times 10^{-7}$ | $2.65 \times 10^{-1}$ | $3.66 \times 10^{-2}$ |
| ***BSN*** | 3 | 49554477 | SGIT | ncod | $2.15 \times 10^{-9}$ | $2.15 \times 10^{-9}$ | **$2.13 \times 10^{-5}$** | **$8.04 \times 10^{-12}$** |
| *C15orf39* | 15 | 75195643 | SGIT | nsyn | $1.06 \times 10^{-7}$ | $1.06 \times 10^{-7}$ | $2.80 \times 10^{-2}$ | $2.48 \times 10^{-1}$ |
| | | | SGIT | cod | $1.12 \times 10^{-7}$ | $1.12 \times 10^{-7}$ | $1.13 \times 10^{-2}$ | $3.80 \times 10^{-1}$ |
| ***C8orf34*** | 8 | 68330955 | CBP | ncod | $3.38 \times 10^{-7}$ | $3.38 \times 10^{-7}$ | **$3.01 \times 10^{-4}$** | $1.98 \times 10^{-1}$ |
| ***CHST3*** | 10 | 71964395 | IDD | ncod | $1.74 \times 10^{-7}$ | $1.74 \times 10^{-7}$ | **$3.21 \times 10^{-17}$** | **$2.09 \times 10^{-11}$** |
| ***COL11A1*** | 1 | 102876467 | SGIT | ncod | $4.76 \times 10^{-7}$ | $4.76 \times 10^{-7}$ | **$2.05 \times 10^{-9}$** | **$1.34 \times 10^{-4}$** |
| ***DCC*** | 18 | 52340197 | SGIT | ncod | $4.65 \times 10^{-9}$ | $4.65 \times 10^{-9}$ | **$8.50 \times 10^{-13}$** | **$1.60 \times 10^{-13}$** |
| | | | CBP | ncod | $1.27 \times 10^{-8}$ | $1.27 \times 10^{-8}$ | | |
| *EXD3* | 9 | 137306896 | CBP | ncod | $3.90 \times 10^{-9}$ | $3.90 \times 10^{-9}$ | $5.76 \times 10^{-1}$ | $1.78 \times 10^{-3}$ |
| | | | SGIT | ncod | $6.47 \times 10^{-7}$ | $6.47 \times 10^{-7}$ | | |
| *FAM172A* | 5 | 93617725 | SGIT | ncod | $1.06 \times 10^{-6}$ | $1.06 \times 10^{-6}$ | $3.11 \times 10^{-1}$ | $1.13 \times 10^{-2}$ |
| *GABRB2* | 5 | 161288429 | CBP | ncod | $2.04 \times 10^{-6}$ | $2.04 \times 10^{-6}$ | $6.79 \times 10^{-2}$ | $5.64 \times 10^{-3}$ |
| ***ILRUN*** | 6 | 34587288 | SGIT | ncod | $1.16 \times 10^{-8}$ | $1.16 \times 10^{-8}$ | **$2.21 \times 10^{-5}$** | **$2.97 \times 10^{-5}$** |
| | | | CBP | ncod | $7.69 \times 10^{-7}$ | $7.69 \times 10^{-7}$ | | |
| *IP6K3* | 6 | 33721662 | SGIT | ncod | $4.10 \times 10^{-7}$ | $2.31 \times 10^{-7}$ | $6.25 \times 10^{-2}$ | $1.52 \times 10^{-2}$ |
| ***JADE2*** | 5 | 134524312 | SGIT | ncod | $4.40 \times 10^{-7}$ | $4.40 \times 10^{-7}$ | **$3.68 \times 10^{-4}$** | $1.20 \times 10^{-2}$ |
| *JAKMIP3* | 10 | 132036336 | CBP | ncod | $2.44 \times 10^{-6}$ | $2.44 \times 10^{-6}$ | $3.70 \times 10^{-3}$ | $3.23 \times 10^{-1}$ |
| *KNTC1* | 12 | 122527246 | CBP | ncod | $6.29 \times 10^{-8}$ | $6.29 \times 10^{-8}$ | $6.87 \times 10^{-2}$ | $4.54 \times 10^{-3}$ |
| | | | SGIT | ncod | $2.16 \times 10^{-6}$ | $2.16 \times 10^{-6}$ | | |
| *LANCL1* | 2 | 210431249 | SGIT | ncod | $1.48 \times 10^{-9}$ | $1.48 \times 10^{-9}$ | $8.56 \times 10^{-1}$ | $2.91 \times 10^{-1}$ |
| ***MAML3*** | 4 | 139716753 | SGIT | ncod | $1.65 \times 10^{-8}$ | $1.65 \times 10^{-8}$ | $5.00 \times 10^{-2}$ | **$1.49 \times 10^{-4}$** |
| | | | CBP | ncod | $1.39 \times 10^{-6}$ | $1.64 \times 10^{-6}$ | | |
| ***MIPOL1*** | 14 | 37197894 | CBP | ncod | $8.41 \times 10^{-8}$ | $8.41 \times 10^{-8}$ | **$8.64 \times 10^{-6}$** | **$9.11 \times 10^{-4}$** |
| | | | SGIT | ncod | $5.54 \times 10^{-7}$ | $5.54 \times 10^{-7}$ | | |
| ***MLLT10*** | 10 | 21524646 | SGIT | ncod | $1.30 \times 10^{-6}$ | $1.30 \times 10^{-6}$ | $8.10 \times 10^{-2}$ | **$9.57 \times 10^{-5}$** |
| *MRPS21* | 1 | 150293861 | CBP | ncod | $3.48 \times 10^{-7}$ | $3.48 \times 10^{-7}$ | $2.25 \times 10^{-1}$ | $2.67 \times 10^{-1}$ |
| *NUDT12* | 5 | 103548855 | CBP | ncod | $3.07 \times 10^{-7}$ | $3.13 \times 10^{-7}$ | $1.46 \times 10^{-1}$ | $3.07 \times 10^{-1}$ |
| ***PTPRC*** | 1 | 198638457 | SGIT | ncod | $2.28 \times 10^{-6}$ | $2.28 \times 10^{-6}$ | **$1.83 \times 10^{-5}$** | **$4.91 \times 10^{-4}$** |
| ***RERG*** | 12 | 15107783 | SGIT | ncod | $4.39 \times 10^{-7}$ | $4.39 \times 10^{-7}$ | **$7.26 \times 10^{-4}$** | **$9.67 \times 10^{-6}$** |
| ***RHOA*** | 3 | 49359139 | CBP | ncod | $1.05 \times 10^{-8}$ | $1.05 \times 10^{-8}$ | **$5.17 \times 10^{-5}$** | **$4.68 \times 10^{-7}$** |
| ***SMAD3*** | 15 | 67063763 | SGIT | ncod | $4.55 \times 10^{-7}$ | $4.55 \times 10^{-7}$ | **$5.81 \times 10^{-17}$** | **$4.97 \times 10^{-4}$** |
| *SON* | 21 | 33543038 | SGIT | ncod | $1.57 \times 10^{-6}$ | $1.57 \times 10^{-6}$ | $3.78 \times 10^{-3}$ | $2.72 \times 10^{-1}$ |
| ***SOX5*** | 12 | 23529504 | CBP | ncod | $7.50 \times 10^{-13}$ | $7.50 \times 10^{-13}$ | **$2.3810^{-25}$** | **$1.33 \times 10^{-18}$** |
| | | | SGIT | ncod | $1.77 \times 10^{-11}$ | $1.77 \times 10^{-11}$ | | |
| ***SPOCK2*** | 10 | 72059034 | SGIT | ncod | $8.10 \times 10^{-13}$ | $8.10 \times 10^{-13}$ | **$6.91 \times 10^{-11}$** | **$1.04 \times 10^{-6}$** |
| | | | CBP | ncod | $2.69 \times 10^{-10}$ | $2.69 \times 10^{-10}$ | | |
| *TCF4* | 18 | 55222185 | SGIT | ncod | $8.89 \times 10^{-7}$ | $1.68 \times 10^{-6}$ | $4.98 \times 10^{-2}$ | $2.45 \times 10^{-2}$ |
| *TMEM59L* | 19 | 18607430 | SGIT | ncod | $6.82 \times 10^{-7}$ | $6.82 \times 10^{-7}$ | $1.82 \times 10^{-2}$ | $7.28 \times 10^{-3}$ |
| *TRAPPC2L* | 16 | 88856220 | CBP | ncod | $2.04 \times 10^{-7}$ | $2.04 \times 10^{-7}$ | $2.96 \times 10^{-1}$ | $1.86 \times 10^{-2}$ |

**Figure 2.**
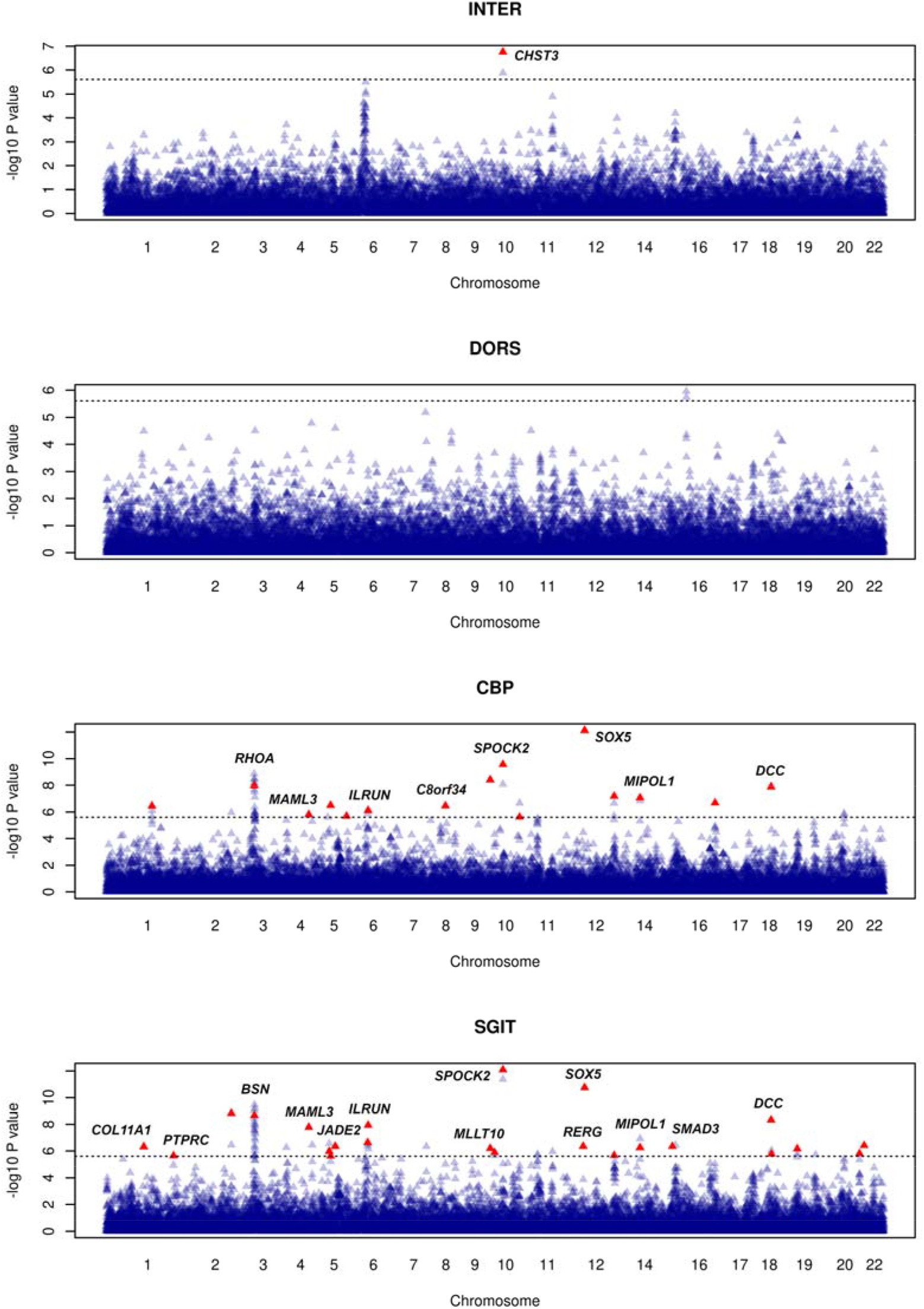
Manhattan plots for gene-based association analysis using noncoding variants. Blue and red triangles indicate -log10(P value) for analyzed genes before and after conditional analysis, respectively. Replicated genes are labeled. Dotted line indicates the significance level (2.5 × 10^-6^).

For 16 replicated genes, we compared the p-values obtained on SGIT and original BP-related phenotypes (Table 2).

**Table 2.** Results of association analysis of original traits for genes identified on SGIT.

| Gene | Chr | Position | SGIT | Gene-based p-value |  |  |
| --- | --- | --- | --- | --- | --- | --- |
|  |  |  |  | CBP | IDD | Dorsalgia |
| *COL11A1* | 1 | 102876467 | $4.76 \times 10^{-7}$ | $7.49 \times 10^{-4}$ | $5.14 \times 10^{-4}$ | $3.21 \times 10^{-5}$ |
| *PTPRC* | 1 | 198638457 | $2.28 \times 10^{-6}$ | $9.70 \times 10^{-4}$ | $2.32 \times 10^{-3}$ | $2.82 \times 10^{-2}$ |
| *BSN* | 3 | 49554477 | $2.15 \times 10^{-9}$ | $8.42 \times 10^{-9}$ | $9.85 \times 10^{-2}$ | $1.83 \times 10^{-2}$ |
| *MAML3* | 4 | 139716753 | $1.65 \times 10^{-8}$ | $1.39 \times 10^{-6}$ | $5.45 \times 10^{-2}$ | $5.32 \times 10^{-4}$ |
| *JADE2* | 5 | 134524312 | $4.40 \times 10^{-7}$ | $3.42 \times 10^{-4}$ | $1.74 \times 10^{-2}$ | $2.50 \times 10^{-5}$ |
| *ILRUN* | 6 | 34587288 | $1.16 \times 10^{-8}$ | $7.69 \times 10^{-7}$ | $1.14 \times 10^{-2}$ | $9.05 \times 10^{-4}$ |
| *MLLT10* | 10 | 21524646 | $1.30 \times 10^{-6}$ | $4.82 \times 10^{-5}$ | $3.40 \times 10^{-1}$ | $1.33 \times 10^{-3}$ |
| *SPOCK2* | 10 | 72059034 | $8.10 \times 10^{-13}$ | $2.69 \times 10^{-10}$ | $1.32 \times 10^{-6}$ | $8.66 \times 10^{-3}$ |
| *RERG* | 12 | 15107783 | $4.39 \times 10^{-7}$ | $3.45 \times 10^{-5}$ | $6.56 \times 10^{-2}$ | $5.15 \times 10^{-4}$ |
| *SOX5* | 12 | 23529504 | $1.77 \times 10^{-11}$ | $7.50 \times 10^{-13}$ | $9.97 \times 10^{-1}$ | $1.23 \times 10^{-1}$ |
| *MIPOL1* | 14 | 37197894 | $5.54 \times 10^{-7}$ | $8.41 \times 10^{-8}$ | $4.12 \times 10^{-1}$ | $3.47 \times 10^{-2}$ |
| *SMAD3* | 15 | 67063763 | $4.55 \times 10^{-7}$ | $7.26 \times 10^{-5}$ | $6.82 \times 10^{-4}$ | $1.56 \times 10^{-2}$ |
| *DCC* | 18 | 52340197 | $4.65 \times 10^{-9}$ | $1.27 \times 10^{-8}$ | $9.97 \times 10^{-1}$ | $2.11 \times 10^{-3}$ |

## 4. Discussion

We have performed gene-based analysis of three BP-related phenotypes and their multi-trait combination using the UK Biobank data. We identified and replicated 16 genes, 13 of which were detected using multi-trait phenotype SGIT. Seven genes identified using SGIT were undetected using the original BP-related phenotypes. In recent years, several GWAS of BP-related phenotypes have been performed using the UK Biobank data [5,6]. However, only a few independent loci were identified and replicated [25]. The success of our study in detecting BP-related genes can be explained by the new methodology used for analysis.

In our study, two approaches have been newly applied for BP-related phenotypes: gene-based and multi-trait association analyses. The gene-based association analysis of BP has been previously performed only for the analysis of rare variants from exome sequenced data [11,26] but not imputed genotypes.

Additionally, a multi-trait approach was used to solve the following problem. According to a contemporary model, there is a complex interplay between biological, psychological, and social factors that contribute to back pain [27]. The identification of novel genetic loci is severely hampered by the absence of a consensus definition of BP. In order to overcome these obstacles and improve our comprehension of the genetic impact on BP, we employed a novel approach. Three distinct phenotypes related to back pain were taken into consideration: two were based on EHR-based codes, and one was self-reported. Dorsalgia and IDD are two EHR-based back pain-related phenotypes that are different from self-reported CBP in that they are based on longitudinal health care records, frequently spanning many years, rather than point prevalence from a single survey. They reflect back pain that is severe enough to warrant seeking medical attention. Nevertheless, due to the roughly 90% genetic correlations among these phenotypes, the genetic nature of all of them is common. However, it does not mean that all three BP-related phenotypes will manifest in the majority of the patients. The discordance between phenotypes is explained by the relatively low heritability (about 50%) of each phenotype. Due to this, only some patients manifested more than one BP-related phenotype (Supplementary Figure S1).

We explored shared pathways among different pain phenotypes using multi-trait methodologies, which can help reduce heterogeneity [8,12]. We used SHAHER methodology [12] for multi-trait analysis. SHAHER constructs a multi-trait phenotype as a linear combination of original traits, where the coefficients are estimated to maximize the heritability of the multi-trait phenotype. As a result, we identified seven genes associated with SGIT, which did not show a significant association with the original traits. Additional six genes were associated with both SGIT and CBP. For the majority of genes associated with SGIT, the p-values obtained on the original BP-related traits were rather low (Table 2). These results confirm the effectiveness of the chosen strategy.

Earlier, we applied both methodologies to analyze the associations between BP and rare genetic variants using exome sequenced data [11]. We identified a new *FSCN3* gene associated with BP-related multi-trait phenotype due to its loss of function (LOF) variants. These variants changing protein structure can be identified when exome sequenced data is used. Imputed genotypes have better coverage of non-coding variants. All genes identified and replicated in the current study showed an association due to non-coding variants (Table 1). Therefore, association analysis using imputed and sequenced genotypes provided different knowledge about the genetic architecture of BP.

The highest association signals were detected for two genes, *SOX5* (p = 1.8×10^-11^ for SGIT and p = 7.5×10^-13^ for CBP) and *SPOCK2* (p = 8.1×10^-13^ for SGIT and p = 2.7×10^-10^ for CBP). The *SOX5* gene codes transcription factors that play essential roles in chondrocyte differentiation [28]; this gene has been previously detected in [4] and by our group as associated with BP-related phenotypes [4-6]. The association of *SPOCK2* was described in [5]. It is interesting that *SPOCK2* is located close to the *CHST3* gene which codes for an enzyme that catalyzes proteoglycan sulfating and has been identified as a susceptibility gene for lumbar disc degeneration [29]. The association of *CHST3* with IDD and dorsalgia was described originally in a study of Asian participants [4]. Moreover, this gene is considered as most probably causal for back pain in this region (cumulative evidence L2G scores for *CHST3* and *SPOCK2* are 0.81 and 0.23, respectively, https://genetics.opentargets.org/study-locus/NEALE2_6159_4/10_72001257_A_G [30]. We believe that a significant result for the *SPOCK2* gene indicates an inability of association analysis to resolve the causal gene in the region, especially in cases with a strong LD block. In such instances, utilizing GWAS results from other ancestries has the potential to provide assistance.

For the additional six genes identified: *BSN, DCC, ILRUN, SMAD3, COL11A*, and *C8orf34*, the association with BP-related phenotypes has already been shown [4,6] and our results can be considered as verification of the impact of these genes on BP-related traits.

For the other 7 genes, no direct association with back pain phenotypes has been shown yet. However, three of them were located in known loci, but have not been prioritized: *MIPOL1 / SLC25A21, PTPRC* / *MIR181A1HG, RHOA / BSN* [4]. Current data do not provide enough information to confidently choose a single candidate gene within these loci. For *PTPRC*, the association with inflammatory bowel disease including Crohn’s disease has been shown [31]. The inflammatory conditions have significant genetic correlations with back and multisite chronic pain [32]. *MAML3* has been shown to contribute to several pathways significantly associated with CBP [33]. This gene enables transcription coactivator activity; it is involved in the Notch signaling pathway and positive regulation of transcription by RNA polymerase II. For three further genes: *RERG, JADE2*, and *MLLT10*, association has been found in a multi-trait analysis of endometriosis and multi-site chronic pain [32]. In our study, we observed these genes to be associated with SGIT but not the other phenotypes. As they seem to affect many comorbid pain conditions, this is in line with the multi-trait nature of SGIT. This conclusion is supported by the fact that some genes associated with SGIT are involved in chondrogenesis and inflammatory processes, which are common for BP conditions. In accordance with the GWAS catalog, many genes are associated with the neurological processes (cognitive ability, educational attainment, attention deficit, intelligence, depression, and others), immunological and inflammatory processes (Crohn’s disease, ulcerative colitis, asthma, osteoarthritis, ankylosing spondylitis), and bone mineral density. All these processes can be involved in BP [4,6,34]. However, the exact role of many identified genes in BP is unknown and needs to be clarified.

Our study has several limitations. First, we used only white Europeans because the linear combination coefficients for SGIT cannot be estimated using GWAS of small sample sizes. Therefore, not all our findings and conclusions could be applied to other ethnic groups. Secondly, we replicated the association signals using the freely available FinnGen summary statistics for dorsalgia and IDD. The summary statistics for CBP were absent in this data set and were not included in replication. We replicated 16 of the 32 genes that passed conditional analysis. It is possible that some of the non-replicated genes will be replicated later, when summary statistics obtained from other large projects will be accessible for replication.

## 5. Conclusions

We conducted a wide gene-based association analysis of BP-related phenotypes using imputed genotypes from a UK Biobank project and applying the technique of multi-trait analysis. We identified and replicated 16 genes significantly associated with BP-related traits. Thirteen genes were detected on multi-trait phenotype that is in accordance with high genetic correlation between BP-related traits. Some genes have been previously described as associated with BP or with the genetically correlated traits or as included in pathways associated with BP.

## Data Availability

The results of the gene-based analysis are available in the ZENODO database: https://doi.org/10.5281/zenodo.8118630.
The original data are available at:
https://www.ukbiobank.ac.uk/
https://www.finngen.fi/

## Author Contributions

Y.A.T. and T.I.A. conceived and oversaw the study; they contributed to the design of the study and interpretation of the results. N.M.B. and I.V.Z. performed a gene-based association analysis. E.E.E. performed genome-wide association analysis, and SHAHER analysis. N.M.B. performed a conditional association analysis. A.V.K., M.F., P.S. and F.W. prepared material. G.R.S. wrote the source code to optimize computations. T.I.A. wrote the first draft of the manuscript. All co-authors discussed the results and contributed to preparing the final version of the manuscript. All authors have read and agreed to the published version of the manuscript.

## Funding

The work of Y.A.T., T.I.A., E.E.E., and N.M.B. was supported by the Russian Science Foundation (RSF) grant No. 22-15-20037 and the Government of the Novosibirsk region.

## Institutional Review Board Statement

Not applicable

## Informed Consent Statement

Not applicable

## Data Availability Statement

The results of the gene-based analysis are available in the ZENODO database: https://doi.org/10.5281/zenodo.8118630.

## Acknowledgments

This research has been conducted using the UK Biobank Resource under Applications #18219 and #59345. We want to acknowledge the participants and investigators of the FinnGen study.

## Conflicts of Interest

The authors declare no conflict of interest. The funders had no role in the design of the study; in the collection, analyses, or interpretation of data; in the writing of the manuscript; or in the decision to publish the results.

## Notes

### Competing Interest Statement

The authors have declared no competing interest.

## References

1. Hoy, D.; March, L.; Brooks, P.; Blyth, F.; Woolf, A.; Bain, C.; Williams, G.; Smith, E.; Vos, T.; Barendregt, J.; et al. The global burden of low back pain: estimates from the Global Burden of Disease 2010 study. Ann Rheum Dis 2014, 73, 968–974, doi:10.1136/annrheumdis-2013-204428.

2. Manchikanti, L.; Singh, V.; Falco, F.J.; Benyamin, R.M.; Hirsch, J.A. Epidemiology of low back pain in adults. Neuromodulation 2014, 17 Suppl 2, 3–10, doi:10.1111/ner.12018.

3. Hartvigsen, J.; Nielsen, J.; Kyvik, K.O.; Fejer, R.; Vach, W.; Iachine, I.; Leboeuf-Yde, C. Heritability of spinal pain and consequences of spinal pain: a comprehensive genetic epidemiologic analysis using a population-based sample of 15,328 twins ages 20-71 years. Arthritis Rheum 2009, 61, 1343–1351, doi:10.1002/art.24607.

4. Bjornsdottir, G.; Stefansdottir, L.; Thorleifsson, G.; Sulem, P.; Norland, K.; Ferkingstad, E.; Oddsson, A.; Zink, F.; Lund, S.H.; Nawaz, M.S.; et al. Rare SLC13A1 variants associate with intervertebral disc disorder highlighting role of sulfate in disc pathology. Nat Commun 2022, 13, 634, doi:10.1038/s41467-022-28167-1.

5. Freidin, M.B.; Tsepilov, Y.A.; Palmer, M.; Karssen, L.C.; Suri, P.; Aulchenko, Y.S.; Williams, F.M.K.; Group, C.M.W. Insight into the genetic architecture of back pain and its risk factors from a study of 509,000 individuals. Pain 2019, 160, 1361–1373, doi:10.1097/j.pain.0000000000001514.

6. Suri, P.; Palmer, M.R.; Tsepilov, Y.A.; Freidin, M.B.; Boer, C.G.; Yau, M.S.; Evans, D.S.; Gelemanovic, A.; Bartz, T.M.; Nethander, M.; et al. Genome-wide meta-analysis of 158,000 individuals of European ancestry identifies three loci associated with chronic back pain. PLoS Genet 2018, 14, e1007601, doi:10.1371/journal.pgen.1007601.

7. Suri, P.; Stanaway, I.B.; Zhang, Y.; Freidin, M.B.; Tsepilov, Y.A.; Carrell, D.S.; Williams, F.M.K.; Aulchenko, Y.S.; Hakonarson, H.; Namjou, B.; et al. Genome-wide association studies of low back pain and lumbar spinal disorders using electronic health record data identify a locus associated with lumbar spinal stenosis. Pain 2021, 162, 2263–2272, doi:10.1097/j.pain.0000000000002221.

8. Tsepilov, Y.A.; Freidin, M.B.; Shadrina, A.S.; Sharapov, S.Z.; Elgaeva, E.E.; Zundert, J.V.; Karssen Lcapital Es, C.; Suri, P.; Williams, F.M.K.; Aulchenko, Y.S. Analysis of genetically independent phenotypes identifies shared genetic factors associated with chronic musculoskeletal pain conditions. Commun Biol 2020, 3, 329, doi:10.1038/s42003-020-1051-9.

9. Polderman, T.J.; Benyamin, B.; de Leeuw, C.A.; Sullivan, P.F.; van Bochoven, A.; Visscher, P.M.; Posthuma, D. Meta-analysis of the heritability of human traits based on fifty years of twin studies. Nat Genet 2015, 47, 702–709, doi:10.1038/ng.3285.

10. Bjornsdottir, G.; Benonisdottir, S.; Sveinbjornsson, G.; Styrkarsdottir, U.; Thorleifsson, G.; Walters, G.B.; Bjornsson, A.; Olafsson, I.H.; Ulfarsson, E.; Vikingsson, A.; et al. Sequence variant at 8q24.21 associates with sciatica caused by lumbar disc herniation. Nat Commun 2017, 8, 14265, doi:10.1038/ncomms14265.

11. Zorkoltseva, I.V.; Elgaeva, E.E.; Belonogova, N.M.; Kirichenko, A.V.; Svishcheva, G.R.; Freidin, M.B.; Williams, F.M.K.; Suri, P.; Tsepilov, Y.A.; Axenovich, T.I. Multi-Trait Exome-Wide Association Study of Back Pain-Related Phenotypes. Genes (Basel) 2023, 14, doi:10.3390/genes14101962.

12. Svishcheva, G.R.; Tiys, E.S.; Elgaeva, E.E.; Feoktistova, S.G.; Timmers, P.; Sharapov, S.Z.; Axenovich, T.I.; Tsepilov, Y.A. A novel framework for analysis of the shared genetic background of correlated traits. Genes (Basel) 2022, 13, 1694, doi:10.3390/genes13101694.

13. Li, B.; Leal, S.M. Methods for detecting associations with rare variants for common diseases: application to analysis of sequence data. Am J Hum Genet 2008, 83, 311–321, doi:10.1016/j.ajhg.2008.06.024.

14. Bycroft, C.; Freeman, C.; Petkova, D.; Band, G.; Elliott, L.T.; Sharp, K.; Motyer, A.; Vukcevic, D.; Delaneau, O.; O’Connell, J.; et al. The UK Biobank resource with deep phenotyping and genomic data. Nature 2018, 562, 203–209, doi:10.1038/s41586-018-0579-z.

15. McCarthy, S.; Das, S.; Kretzschmar, W.; Delaneau, O.; Wood, A.R.; Teumer, A.; Kang, H.M.; Fuchsberger, C.; Danecek, P.; Sharp, K.; et al. A reference panel of 64,976 haplotypes for genotype imputation. Nat Genet 2016, 48, 1279–1283, doi:10.1038/ng.3643.

16. Jiang, L.; Zheng, Z.; Fang, H.; Yang, J. A generalized linear mixed model association tool for biobank-scale data. Nat Genet 2021, 53, 1616–1621, doi:10.1038/s41588-021-00954-4.

17. McLaren, W.; Gil, L.; Hunt, S.E.; Riat, H.S.; Ritchie, G.R.; Thormann, A.; Flicek, P.; Cunningham, F. The Ensembl Variant Effect Predictor. Genome Biol 2016, 17, 122, doi:10.1186/s13059-016-0974-4.

18. Lee, S.; Wu, M.C.; Lin, X. Optimal tests for rare variant effects in sequencing association studies. Biostatistics 2012, 13, 762–775, doi:10.1093/biostatistics/kxs014.

19. Wang, K.; Abbott, D. A principal components regression approach to multilocus genetic association studies. Genet Epidemiol 2008, 32, 108–118, doi:10.1002/gepi.20266.

20. Liu, Y.; Chen, S.; Li, Z.; Morrison, A.C.; Boerwinkle, E.; Lin, X. ACAT: A Fast and Powerful p Value Combination Method for Rare-Variant Analysis in Sequencing Studies. Am J Hum Genet 2019, 104, 410–421, doi:10.1016/j.ajhg.2019.01.002.

21. Svishcheva, G.R.; Belonogova, N.M.; Zorkoltseva, I.V.; Kirichenko, A.V.; Axenovich, T.I. Gene-based association tests using GWAS summary statistics. Bioinformatics 2019, 35, 3701–3708, doi:10.1093/bioinformatics/btz172.

22. Yang, J.; Ferreira, T.; Morris, A.P.; Medland, S.E.; Genetic Investigation of, A.T.C.; Replication, D.I.G.; Meta-analysis, C.; Madden, P.A.; Heath, A.C.; Martin, N.G.; et al. Conditional and joint multiple-SNP analysis of GWAS summary statistics identifies additional variants influencing complex traits. Nat Genet 2012, 44, 369-375, S361-363, doi:10.1038/ng.2213.

23. Yang, J.; Lee, S.H.; Goddard, M.E.; Visscher, P.M. GCTA: a tool for genome-wide complex trait analysis. Am J Hum Genet 2011, 88, 76–82, doi:10.1016/j.ajhg.2010.11.011.

24. Kurki, M.I.; Karjalainen, J.; Palta, P.; Sipila, T.P.; Kristiansson, K.; Donner, K.M.; Reeve, M.P.; Laivuori, H.; Aavikko, M.; Kaunisto, M.A.; et al. FinnGen provides genetic insights from a well-phenotyped isolated population. Nature 2023, 613, 508–518, doi:10.1038/s41586-022-05473-8.

25. Li, S.; Brimmers, A.; van Boekel, R.L.M.; Vissers, K.C.P.; Coenen, M.J.H. A systematic review of genome-wide association studies for pain, nociception, neuropathy, and pain treatment responses. Pain 2023, 164, 1891–1911, doi:10.1097/j.pain.0000000000002910.

26. Ao, X.; Parisien, M.; Zidan, M.; Grant, A.V.; Martinsen, A.E.; Winsvold, B.S.; Diatchenko, L. Rare variant analyses in large-scale cohorts identified SLC13A1 associated with chronic pain. Pain 2023, 164, 1841–1851, doi:10.1097/j.pain.0000000000002882.

27. Gatchel, R.J.; Mayer, T.G.; Choi, Y.; Chou, R. Validation of a consensus-based minimal clinically important difference (MCID) threshold using an objective functional external anchor. Spine J 2013, 13, 889–893, doi:10.1016/j.spinee.2013.02.015.

28. Yang, Z.; Ren, Z.; She, R.; Ao, J.; Wa, Q.; Sun, Z.; Li, B.; Tian, X. miR-23a-3p regulated by LncRNA SNHG5 suppresses the chondrogenic differentiation of human adipose-derived stem cells via targeting SOX6/SOX5. Cell Tissue Res 2021, 383, 723–733, doi:10.1007/s00441-020-03289-4.

29. Song, Y.Q.; Karasugi, T.; Cheung, K.M.; Chiba, K.; Ho, D.W.; Miyake, A.; Kao, P.Y.; Sze, K.L.; Yee, A.; Takahashi, A.; et al. Lumbar disc degeneration is linked to a carbohydrate sulfotransferase 3 variant. J Clin Invest 2013, 123, 4909–4917, doi:10.1172/JCI69277.

30. Mountjoy, E.; Schmidt, E.M.; Carmona, M.; Schwartzentruber, J.; Peat, G.; Miranda, A.; Fumis, L.; Hayhurst, J.; Buniello, A.; Karim, M.A.; et al. An open approach to systematically prioritize causal variants and genes at all published human GWAS trait-associated loci. Nat Genet 2021, 53, 1527–1533, doi:10.1038/s41588-021-00945-5.

31. Liu, J.Z.; van Sommeren, S.; Huang, H.; Ng, S.C.; Alberts, R.; Takahashi, A.; Ripke, S.; Lee, J.C.; Jostins, L.; Shah, T.; et al. Association analyses identify 38 susceptibility loci for inflammatory bowel disease and highlight shared genetic risk across populations. Nat Genet 2015, 47, 979–986, doi:10.1038/ng.3359.

32. Rahmioglu, N.; Mortlock, S.; Ghiasi, M.; Moller, P.L.; Stefansdottir, L.; Galarneau, G.; Turman, C.; Danning, R.; Law, M.H.; Sapkota, Y.; et al. The genetic basis of endometriosis and comorbidity with other pain and inflammatory conditions. Nat Genet 2023, 55, 423–436, doi:10.1038/s41588-023-01323-z.

33. Bortsov, A.V.; Parisien, M.; Khoury, S.; Martinsen, A.E.; Lie, M.U.; Heuch, I.; Hveem, K.; Zwart, J.A.; Winsvold, B.S.; Diatchenko, L. Brain-specific genes contribute to chronic but not to acute back pain. Pain Rep 2022, 7, e1018, doi:10.1097/PR9.0000000000001018.

34. Naylor, B.; Boag, S.; Gustin, S.M. New evidence for a pain personality? A critical review of the last 120 years of pain and personality. Scand J Pain 2017, 17, 58–67, doi:10.1016/j.sjpain.2017.07.011.

