## Supplementary Materials for "A multi-trait approach identified seven novel genes for back pain-related phenotypes"

**SUPPLEMENTARY INFORMATION**

**Supplementary Methods**

***Chronic back pain (CBP) phenotype definition***

For CBP, cases and controls were defined based on questionnaire responses (data field 3571 “Back pain for 3+ months”). First, participants responded to “Pain type(s) experienced in the last months”, followed by questions inquiring if the specific pain had been present for more than 3 months. Those who reported pain lasting more than 3 months were considered chronic back pain cases, while participants reporting no such pain were considered controls. Individuals who preferred not to answer or reported more than 3 months of pain all over the body were excluded from the study.

***SHAHER analysis***

SHAHER analysis includes two steps. On the first step, the alpha coefficients of the linear combination of the original traits are calculated by the maxSH method. The input data for this method are phenotypic and genetic correlations between the original traits and their SNP-based heritabilities.

Pairwise phenotypic correlations between the three original back pain (BP)-related traits were assessed in a subsample of 307,876 non-relative white individuals with chronic BP (CBP), dorsalgia, and intervertebral disc disorders (IDD) status information. Genetic correlations between the original traits and SNP-based heritabilities were calculated using LD score regression (Bulik-Sullivan et al., 2015) on GWAS results. GWAS summary statistics for the original traits were calculated on imputed genotypes using white British individuals (N = 449,136) by fastGWA-GLMM and were filtered to keep SNVs with imputation quality INFO > 0.8 and MAF > 5×10^-6^. Total number of SNPs was 19,405,718 after filtration. Prior to the genetic correlations and heritability estimation, the summary statistics were reformatted using the munge() function with the default settings from GenomicSEM v0.0.2 R package (Grotzinger et al., 2019).

On the second step of SHAHER, the summary statistics of multi-trait SGIT (shared genetic impact trait) are calculated using the sumCOT method. The input data for sumCOT are the alpha coefficients and the summary statistics obtained for the original traits.

***Phenotypic and genetic correlations, heritability estimates***

Phenotypic correlations of SGIT with CBP, dorsalgia, and IDD were evaluated using summary-level data from imputed genotypes according to the approach proposed by Stephens (2013). We estimated both genetic correlations between SGIT and the original traits and the SGIT SNP-based heritabilitiy utilizing the LD Score regression tool (Bulik-Sullivan et al., 2015) and summary statistics estimated on imputed variant data.

*References*

Bulik-Sullivan BK, Loh PR, Finucane HK, et al. LD Score regression distinguishes confounding from polygenicity in genome-wide association studies. *Nat Genet*. 2015;47(3):291-295.

Grotzinger AD, Rhemtulla M, de Vlaming R, et al. Genomic structural equation modelling provides insights into the multivariate genetic architecture of complex traits. *Nat Hum Behav*. 2019;3(5):513-525.

Stephens M. A unified framework for association analysis with multiple related phenotypes. PLoS One 2013;8(7):e65245.

**Supplementary Figures**

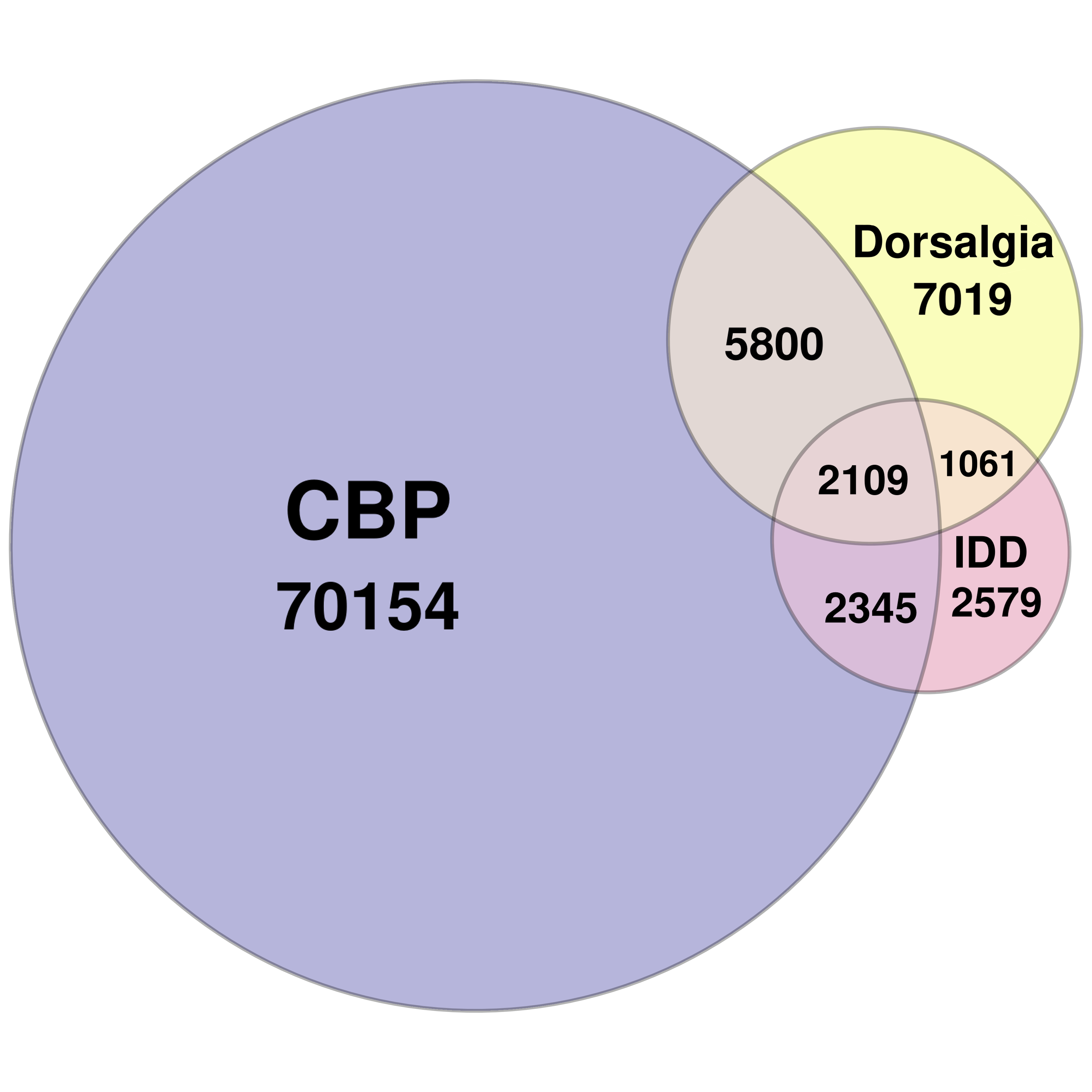

Figure S1. Venn diagram showing the distribution of phenotypes among individuals in 500K data set

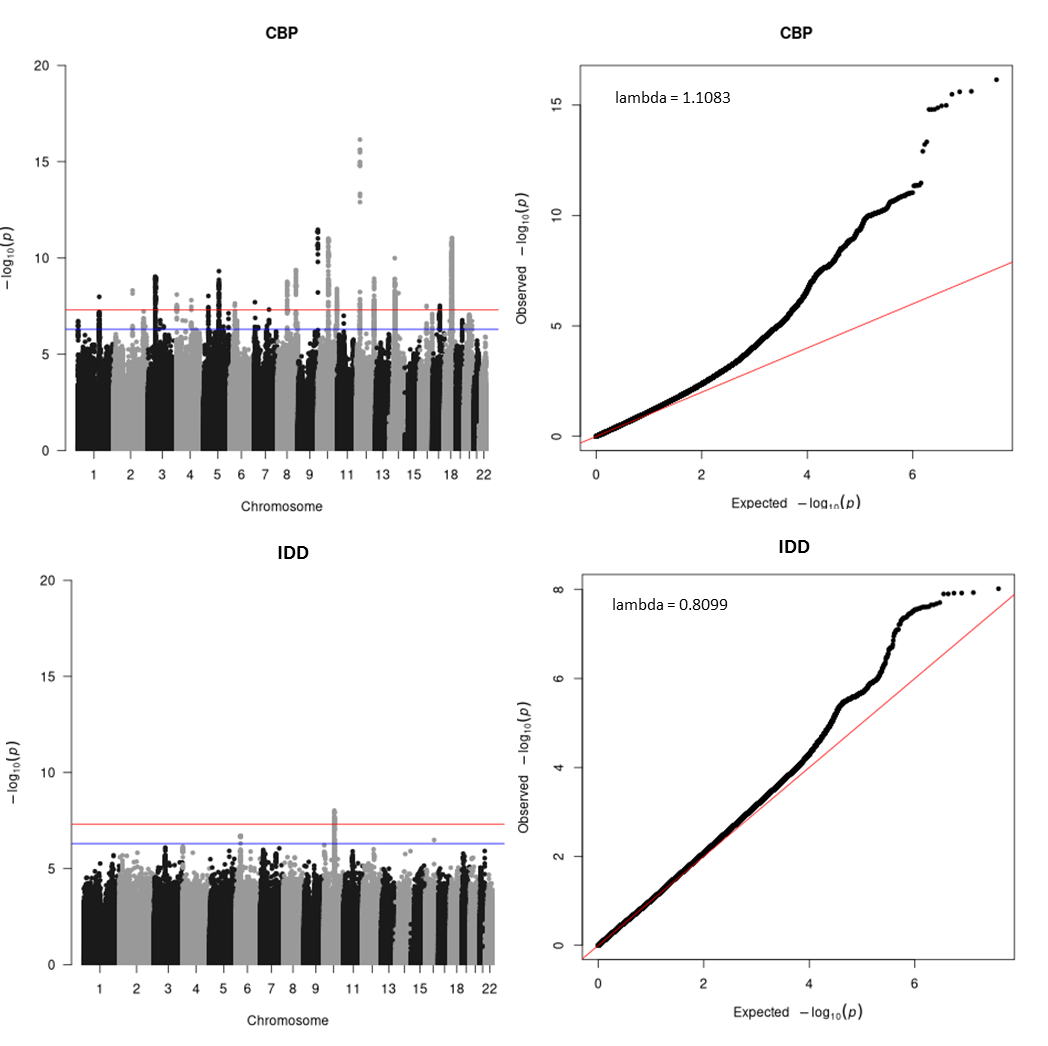

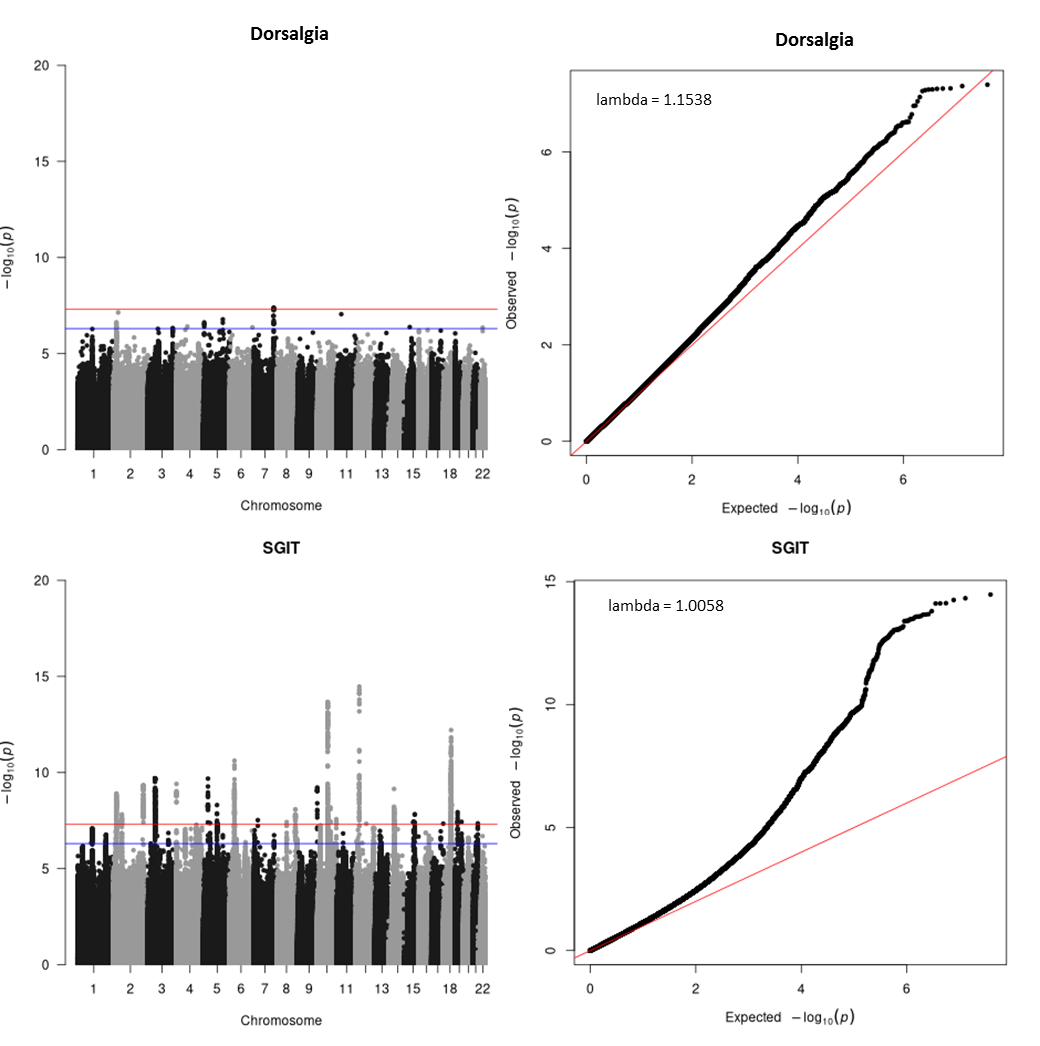

Figure S2. Manhattan (left) and QQ (right) plots for single-point association analyses of different traits using imputed genotypes

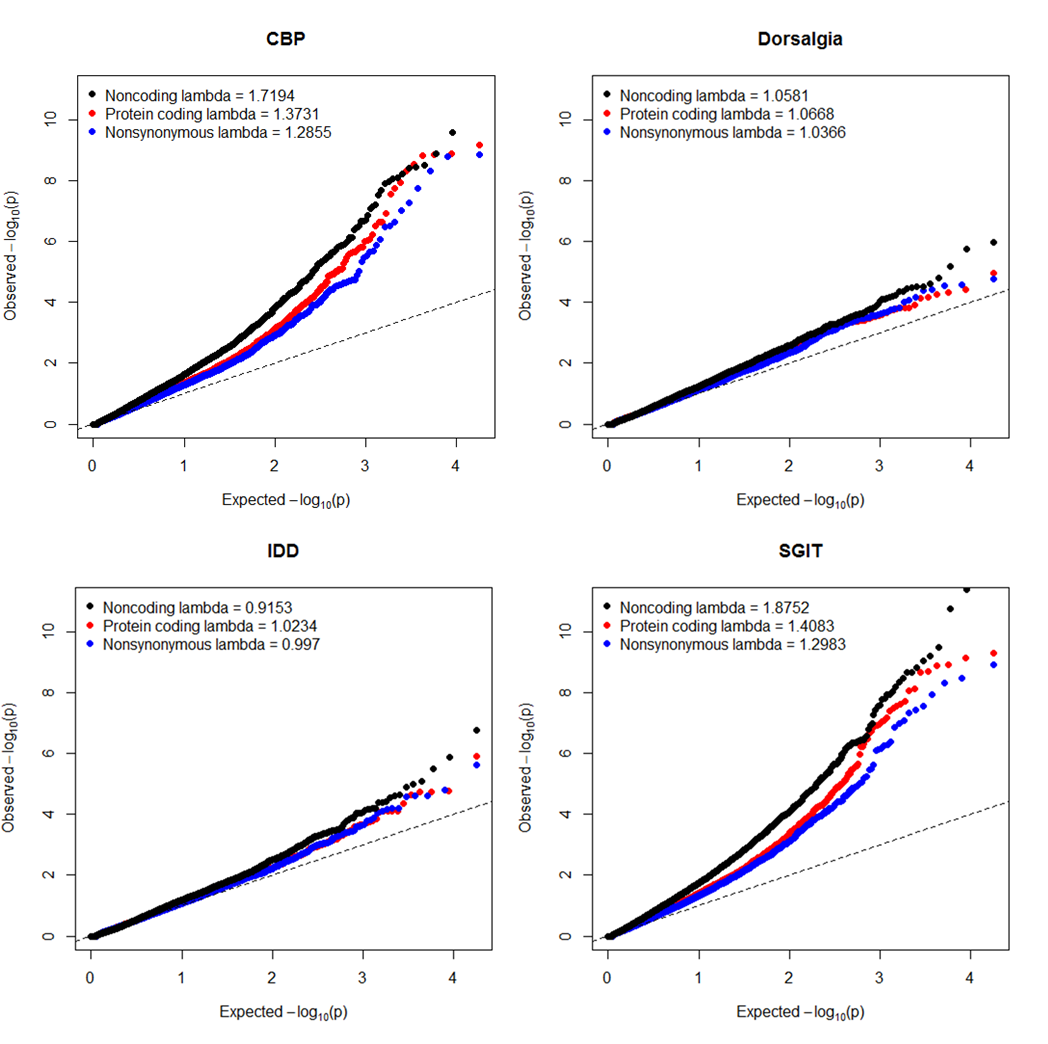

Figure S3. QQ-plots for gene-based analyses of different traits and variant sets using imputed genotypes

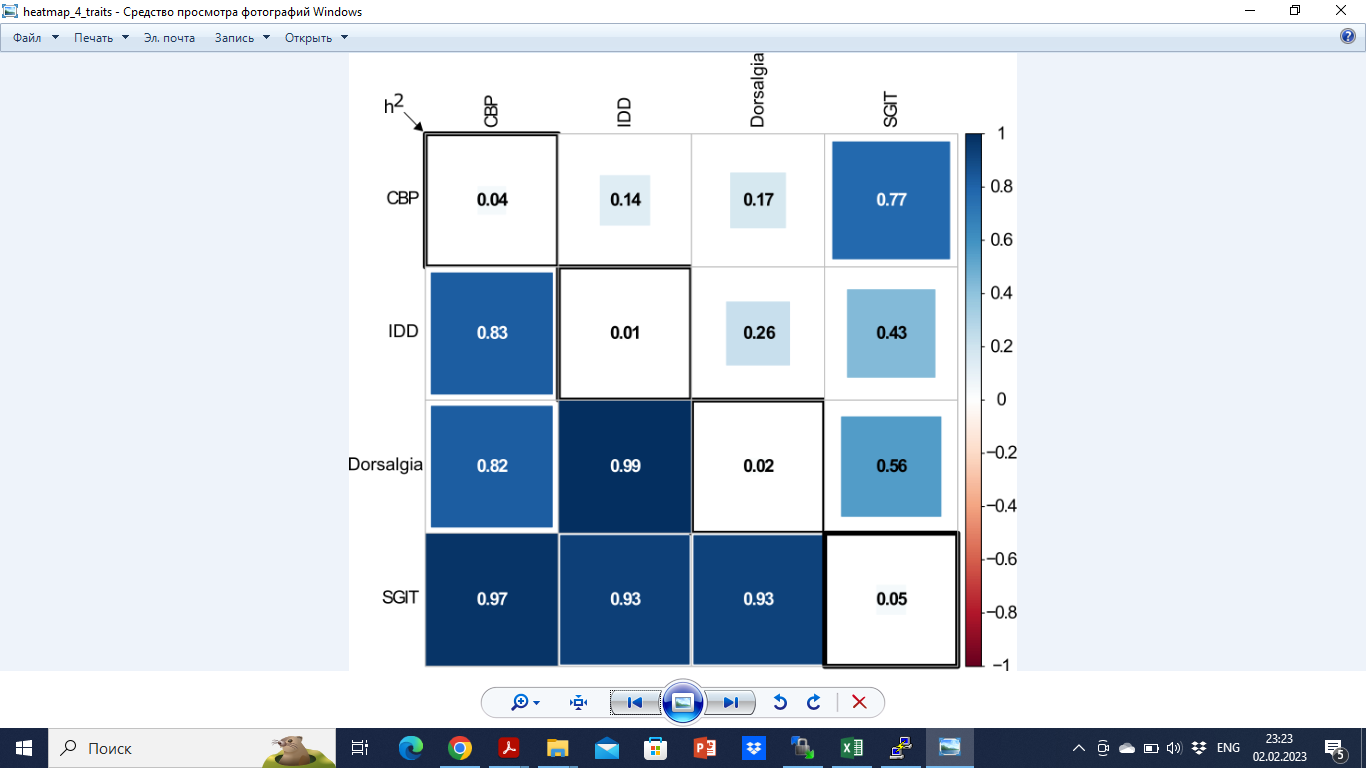

Figure S4. SNP-based heritability (diagonal elements), phenotypic correlations (upper triangular elements) and genetic correlations (lower triangular elements).

**Supplementary Tables**

Table S1. Numbers of variants in different sets

| Set | #SNPs |
| --- | --- |
| Protein coding | 176,949 |
| Protein non-coding | 8,508,956 |
| Nonsynonymous | 106,244 |

Table S2. Results of conditional gene-based association analysis (COJO) of genes showing significant association signals under unconditional analysis (GBA)

| Gene | Chr | Pos | Trait | Set | GBA | COJO |
| --- | --- | --- | --- | --- | --- | --- |
| *COL11A1* | 1 | 102876467 | SGIT | ncod | 4.76×10^-7^ | 4.76×10^-7^ |
| *MRPS21* | 1 | 150293861 | CBP | ncod | 3.48×10^-7^ | 3.48×10^-7^ |
| *PRPF3* | 1 | 150321479 | CBP | ncod | 2.14×10^-6^ | 0.766 |
| *TARS2* | 1 | 150487414 | CBP | ncod | 7.57×10^-7^ | 0.734 |
| *ECM1* | 1 | 150508062 | CBP | cod | 5.87×10^-7^ | 0.486 |
|  |  |  | CBP | nsyn | 3.29×10^-7^ | 0.356 |
| *PTPRC* | 1 | 198638457 | SGIT | ncod | 2.28×10^-6^ | 2.28×10^-6^ |
| *LANCL1* | 2 | 210431249 | CBP | ncod | 1.12×10^-6^ | 0.724 |
|  |  |  | SGIT | ncod | 1.48×10^-9^ | 1.48×10^-9^ |
| *CPS1* | 2 | 210477682 | SGIT | cod | 6.74×10^-8^ | 0.087 |
|  |  |  | SGIT | ncod | 3.47×10^-7^ | 0.647 |
|  |  |  | SGIT | nsyn | 2.73×10^-8^ | 0.087 |
| *GPX1* | 3 | 49357176 | CBP | cod | 1.42×10^-9^ | 0.010 |
|  |  |  | CBP | ncod | 1.35×10^-9^ | 0.027 |
|  |  |  | CBP | nsyn | 1.42×10^-9^ | 0.010 |
|  |  |  | SGIT | cod | 1.26×10^-9^ | 0.100 |
|  |  |  | SGIT | ncod | 8.94×10^-10^ | 0.125 |
|  |  |  | SGIT | nsyn | 1.26×10^-9^ | 0.100 |
| *RHOA* | 3 | 49359139 | CBP | ncod | 1.05×10^-8^ | 1.05×10^-8^ |
|  |  |  | SGIT | ncod | 6.36×10^-9^ | 0.344 |
| *TCTA* | 3 | 49412212 | CBP | ncod | 3.51×10^-9^ | 0.067 |
|  |  |  | SGIT | ncod | 2.11×10^-9^ | 0.212 |
| *AMT* | 3 | 49416778 | CBP | cod | 1.34×10^-9^ | 0.061 |
|  |  |  | CBP | ncod | 5.96×10^-9^ | 0.078 |
|  |  |  | SGIT | cod | 7.15×10^-10^ | 0.042 |
|  |  |  | SGIT | ncod | 3.43×10^-9^ | 0.222 |
| *NICN1* | 3 | 49422333 | CBP | ncod | 2.03×10^-8^ | 0.131 |
|  |  |  | SGIT | ncod | 1.13×10^-8^ | 0.187 |
| *DAG1* | 3 | 49468713 | CBP | cod | 5.00×10^-9^ | 0.767 |
|  |  |  | CBP | ncod | 3.05×10^-8^ | 0.141 |
|  |  |  | SGIT | cod | 7.45×10^-9^ | 0.776 |
|  |  |  | SGIT | ncod | 2.67×10^-8^ | 0.283 |
| *BSN* | 3 | 49554477 | CBP | cod | 1.19×10^-8^ | 0.025 |
|  |  |  | CBP | ncod | 8.42×10^-9^ | 0.018 |
|  |  |  | CBP | nsyn | 3.00×10^-7^ | 0.022 |
|  |  |  | SGIT | cod | 1.32×10^-9^ | 0.011 |
|  |  |  | SGIT | ncod | 2.15×10^-9^ | 2.15×10^-9^ |
|  |  |  | SGIT | nsyn | 4.72×10^-8^ | 0.031 |
| *APEH* | 3 | 49674014 | CBP | cod | 1.85×10^-8^ | 1 |
|  |  |  | CBP | ncod | 3.17×10^-9^ | 0.021 |
|  |  |  | SGIT | cod | 2.20×10^-9^ | 0.056 |
|  |  |  | SGIT | ncod | 3.38×10^-10^ | 0.012 |
| *MST1* | 3 | 49683947 | CBP | cod | 1.52×10^-9^ | 0.058 |
|  |  |  | CBP | ncod | 1.39×10^-6^ | 0.046 |
|  |  |  | CBP | nsyn | 1.84×10^-8^ | 0.913 |
|  |  |  | SGIT | cod | 5.01×10^-10^ | 0.018 |
|  |  |  | SGIT | ncod | 2.73×10^-8^ | 0.172 |
|  |  |  | SGIT | nsyn | 4.74×10^-9^ | 0.399 |
| *RNF123* | 3 | 49689538 | CBP | ncod | 4.22×10^-7^ | 0.026 |
|  |  |  | SGIT | ncod | 1.07×10^-7^ | 0.026 |
| *AMIGO3* | 3 | 49716829 | SGIT | ncod | 2.28×10^-6^ | 0.006 |
| *GMPPB* | 3 | 49716844 | CBP | ncod | 1.28×10^-6^ | 0.021 |
|  |  |  | SGIT | ncod | 5.36×10^-7^ | 0.029 |
| *IP6K1* | 3 | 49724294 | CBP | ncod | 8.97×10^-7^ | 0.036 |
|  |  |  | SGIT | ncod | 3.90×10^-7^ | 0.029 |
| *UBA7* | 3 | 49805209 | SGIT | ncod | 2.12×10^-6^ | 0.049 |
| *TRAIP* | 3 | 49828601 | CBP | cod | 1.55×10^-6^ | 0.002 |
|  |  |  | CBP | ncod | 2.15×10^-6^ | 0.030 |
|  |  |  | SGIT | cod | 2.15×10^-6^ | 0.003 |
|  |  |  | SGIT | ncod | 9.19×10^-7^ | 0.036 |
| *CAMKV* | 3 | 49857988 | CBP | cod | 1.18×10^-7^ | 0.034 |
|  |  |  | CBP | ncod | 7.46×10^-8^ | 0.008 |
|  |  |  | SGIT | cod | 2.05×10^-9^ | 0.011 |
|  |  |  | SGIT | ncod | 6.40×10^-10^ | 0.003 |
| *MST1R* | 3 | 49887002 | CBP | cod | 2.28×10^-6^ | 0.178 |
|  |  |  | CBP | nsyn | 1.31×10^-6^ | 0.114 |
|  |  |  | SGIT | cod | 2.72×10^-8^ | 0.042 |
|  |  |  | SGIT | ncod | 3.82×10^-8^ | 0.045 |
|  |  |  | SGIT | nsyn | 1.20×10^-8^ | 0.031 |
| *MON1A* | 3 | 49907160 | CBP | cod | 1.60×10^-6^ | 0.084 |
|  |  |  | CBP | ncod | 1.06×10^-6^ | 0.076 |
|  |  |  | SGIT | cod | 1.92×10^-8^ | 0.034 |
|  |  |  | SGIT | ncod | 1.57×10^-8^ | 0.034 |
| *RBM6* | 3 | 49940007 | SGIT | cod | 3.66×10^-7^ | 0.002 |
|  |  |  | SGIT | ncod | 5.24×10^-8^ | 0.032 |
| *RBM5* | 3 | 50088919 | SGIT | cod | 1.18×10^-7^ | 0.008 |
|  |  |  | SGIT | ncod | 9.31×10^-9^ | 0.019 |
| *SEMA3F* | 3 | 50155045 | CBP | cod | 1.55×10^-6^ | 0.015 |
|  |  |  | CBP | ncod | 1.51×10^-6^ | 0.007 |
|  |  |  | SGIT | cod | 3.17×10^-8^ | 0.004 |
|  |  |  | SGIT | ncod | 1.57×10^-7^ | 0.005 |
| *GNAT1* | 3 | 50191610 | SGIT | ncod | 2.14×10^-6^ | 0.010 |
| *SPON2* | 4 | 1166932 | SGIT | cod | 2.16×10^-7^ | 0.290 |
|  |  |  | SGIT | ncod | 5.53×10^-7^ | 0.612 |
|  |  |  | SGIT | nsyn | 1.37×10^-7^ | 0.396 |
| *SLC39A8* | 4 | 102251080 | CBP | cod | 2.28×10^-7^ | 0.610 |
|  |  |  | CBP | nsyn | 5.50×10^-8^ | 0.695 |
|  |  |  | SGIT | cod | 4.83×10^-7^ | 0.106 |
|  |  |  | SGIT | nsyn | 4.97×10^-7^ | 1 |
| *MAML3* | 4 | 139716753 | CBP | ncod | 1.39×10^-6^ | 1.64×10^-6^ |
|  |  |  | SGIT | ncod | 1.65×10^-8^ | 1.65×10^-8^ |
| *FNIP2* | 4 | 158769026 | SGIT | ncod | 3.57×10^-7^ | 0.349 |
| *MTX3* | 5 | 79976716 | CBP | ncod | 2.46×10^-6^ | 0.899 |
| *FAM172A* | 5 | 93617725 | SGIT | ncod | 1.06×10^-6^ | 1.06×10^-6^ |
| *POU5F2* | 5 | 93733220 | SGIT | ncod | 2.79×10^-7^ | 0.945 |
| *NUDT12* | 5 | 103548855 | CBP | cod | 2.38×10^-6^ | 0.234 |
|  |  |  | CBP | ncod | 3.07×10^-7^ | 3.13×10^-7^ |
| *JADE2* | 5 | 134524312 | SGIT | ncod | 4.40×10^-7^ | 4.40×10^-7^ |
| *GABRB2* | 5 | 161288429 | CBP | ncod | 2.04×10^-6^ | 2.04×10^-6^ |
| *EGFL8* | 6 | 32164595 | SGIT | nsyn | 7.62×10^-7^ | 0.065 |
| *RNF5* | 6 | 32178405 | IDD | cod | 1.25×10^-6^ | 0.726 |
| *UQCC2* | 6 | 33694293 | CBP | ncod | 1.33×10^-6^ | 0.122 |
| *IP6K3* | 6 | 33721662 | CBP | ncod | 7.47×10^-7^ | 0.297 |
|  |  |  | SGIT | ncod | 4.10×10^-7^ | 2.31×10^-7^ |
| *PACSIN1* | 6 | 34466061 | SGIT | cod | 9.18×10^-8^ | 0.374 |
|  |  |  | SGIT | nsyn | 8.59×10^-8^ | 0.329 |
| *ILRUN* | 6 | 34587288 | CBP | ncod | 7.69×10^-7^ | 7.69×10^-7^ |
|  |  |  | SGIT | ncod | 1.16×10^-8^ | 1.16×10^-8^ |
| *SNRPC* | 6 | 34757505 | SGIT | ncod | 6.95×10^-7^ | 0.829 |
| *UHRF1BP1* | 6 | 34792083 | SGIT | cod | 1.90×10^-7^ | 0.458 |
|  |  |  | SGIT | ncod | 2.75×10^-7^ | 0.878 |
|  |  |  | SGIT | nsyn | 2.40×10^-6^ | 0.375 |
| *MKRN1* | 7 | 140453033 | SGIT | cod | 1.37×10^-7^ | 0.065 |
|  |  |  | SGIT | ncod | 4.54×10^-7^ | 0.037 |
|  |  |  | SGIT | nsyn | 8.17×10^-7^ | 0.131 |
| *KCNH2* | 7 | 150944961 | CBP | nsyn | 2.05×10^-6^ | 1 |
| *C8orf34* | 8 | 68330955 | CBP | ncod | 3.38×10^-7^ | 3.38×10^-7^ |
| *GSDMC* | 8 | 129748196 | CBP | cod | 2.88×10^-8^ | 0.002 |
|  |  |  | SGIT | cod | 8.66×10^-9^ | 0.000 |
| *EXD3* | 9 | 137306896 | CBP | cod | 6.69×10^-10^ | 1 |
|  |  |  | CBP | ncod | 3.90×10^-9^ | 3.90×10^-9^ |
|  |  |  | CBP | nsyn | 1.57×10^-9^ | 1 |
|  |  |  | SGIT | cod | 1.24×10^-7^ | 0.933 |
|  |  |  | SGIT | ncod | 6.47×10^-7^ | 6.47×10^-7^ |
|  |  |  | SGIT | nsyn | 6.84×10^-7^ | 1 |
| *SKIDA1* | 10 | 21513475 | SGIT | ncod | 2.20×10^-6^ | 0.567 |
| *MLLT10* | 10 | 21524646 | SGIT | cod | 3.28×10^-7^ | 0.762 |
|  |  |  | SGIT | ncod | 1.30×10^-6^ | 1.30×10^-6^ |
| *CHST3* | 10 | 71964395 | CBP | ncod | 8.29×10^-9^ | 0.540 |
|  |  |  | IDD | ncod | 1.74×10^-7^ | 1.74×10^-7^ |
|  |  |  | IDD | nsyn | 2.45×10^-6^ | 0.944 |
|  |  |  | SGIT | cod | 2.46×10^-8^ | 0.363 |
|  |  |  | SGIT | ncod | 4.24×10^-12^ | 0.164 |
|  |  |  | SGIT | nsyn | 3.28×10^-9^ | 0.171 |
| *SPOCK2* | 10 | 72059034 | CBP | ncod | 2.69×10^-10^ | 2.69×10^-10^ |
|  |  |  | IDD | ncod | 1.32×10^-6^ | 0.427 |
|  |  |  | SGIT | ncod | 8.10×10^-13^ | 8.10×10^-13^ |
| *ADAM12* | 10 | 126012381 | SGIT | cod | 5.95×10^-7^ | 6.20×10^-7^ |
| *JAKMIP3* | 10 | 132036336 | CBP | cod | 3.09×10^-7^ | 1 |
|  |  |  | CBP | ncod | 2.44×10^-6^ | 2.44×10^-6^ |
|  |  |  | CBP | nsyn | 9.38×10^-8^ | 0.885 |
| *DPYSL4* | 10 | 132186948 | CBP | cod | 9.98×10^-7^ | 0.545 |
|  |  |  | CBP | ncod | 2.09×10^-7^ | 1 |
| *CKAP5* | 11 | 46743048 | SGIT | ncod | 1.71×10^-6^ | 0.001 |
| *LRP4* | 11 | 46856717 | SGIT | ncod | 2.34×10^-6^ | 0.001 |
| *PACS1* | 11 | 66070272 | SGIT | ncod | 1.09×10^-6^ | 0.033 |
| *RERG* | 12 | 15107783 | SGIT | ncod | 4.39×10^-7^ | 4.39×10^-7^ |
| *SOX5* | 12 | 23529504 | CBP | ncod | 7.50×10^-13^ | 7.50×10^-13^ |
|  |  |  | SGIT | ncod | 1.77×10^-11^ | 1.77×10^-11^ |
| *CLIP1* | 12 | 122271432 | CBP | ncod | 1.94×10^-6^ | 0.145 |
| *RSRC2* | 12 | 122503454 | CBP | ncod | 2.13×10^-7^ | 1 |
| *KNTC1* | 12 | 122527246 | CBP | ncod | 6.29×10^-8^ | 6.29×10^-8^ |
|  |  |  | SGIT | ncod | 2.16×10^-6^ | 2.16×10^-6^ |
| *MIPOL1* | 14 | 37197894 | CBP | cod | 2.38×10^-7^ | 1 |
|  |  |  | CBP | ncod | 8.41×10^-8^ | 8.41×10^-8^ |
|  |  |  | CBP | nsyn | 2.31×10^-7^ | 1 |
|  |  |  | SGIT | ncod | 5.54×10^-7^ | 5.54×10^-7^ |
| *FOXA1* | 14 | 37589552 | CBP | cod | 2.27×10^-6^ | 0.087 |
|  |  |  | CBP | ncod | 1.37×10^-7^ | 0.588 |
|  |  |  | CBP | nsyn | 2.18×10^-6^ | 0.071 |
|  |  |  | SGIT | cod | 6.06×10^-7^ | 0.028 |
|  |  |  | SGIT | ncod | 1.17×10^-7^ | 0.262 |
|  |  |  | SGIT | nsyn | 5.74×10^-7^ | 0.026 |
| *SMAD3* | 15 | 67063763 | SGIT | ncod | 4.55×10^-7^ | 4.55×10^-7^ |
| *C15orf39* | 15 | 75195643 | SGIT | cod | 1.12×10^-7^ | 1.12×10^-7^ |
|  |  |  | SGIT | ncod | 3.62×10^-7^ | 0.479 |
|  |  |  | SGIT | nsyn | 1.06×10^-7^ | 1.06×10^-7^ |
| *NTHL1* | 16 | 2039815 | DORS | ncod | 1.08×10^-6^ | 0.123 |
| *TSC2* | 16 | 2047967 | DORS | ncod | 1.82×10^-6^ | 0.529 |
| *TRAPPC2L* | 16 | 88856220 | CBP | ncod | 2.04×10^-7^ | 2.04×10^-7^ |
| *DCC* | 18 | 52340197 | CBP | cod | 2.95×10^-9^ | 0.097 |
|  |  |  | CBP | ncod | 1.27×10^-8^ | 1.27×10^-8^ |
|  |  |  | CBP | nsyn | 4.84×10^-9^ | 0.137 |
|  |  |  | SGIT | cod | 3.88×10^-8^ | 0.788 |
|  |  |  | SGIT | ncod | 4.65×10^-9^ | 4.65×10^-9^ |
|  |  |  | SGIT | nsyn | 3.68×10^-8^ | 0.902 |
| *TCF4* | 18 | 55222185 | SGIT | ncod | 8.89×10^-7^ | 1.68×10^-6^ |
| *CRLF1* | 19 | 18572220 | SGIT | cod | 8.46×10^-8^ | 1 |
|  |  |  | SGIT | ncod | 1.60×10^-6^ | 0.733 |
|  |  |  | SGIT | nsyn | 4.27×10^-7^ | 0.121 |
| *TMEM59L* | 19 | 18607430 | SGIT | ncod | 6.82×10^-7^ | 6.82×10^-7^ |
| *TOMM40* | 19 | 44890569 | CBP | cod | 1.92×10^-6^ | 1 |
| *APOE* | 19 | 44905791 | CBP | cod | 8.89×10^-7^ | 0.697 |
|  |  |  | CBP | nsyn | 8.89×10^-7^ | 0.697 |
| *MYPOP* | 19 | 45890023 | SGIT | ncod | 1.90×10^-6^ | 0.987 |
| *GSS* | 20 | 34928432 | CBP | ncod | 1.62×10^-6^ | 0.515 |
| *MYH7B* | 20 | 34955810 | CBP | ncod | 1.30×10^-6^ | 0.581 |
| *TRPC4AP* | 20 | 35002404 | CBP | cod | 9.51×10^-7^ | 1 |
|  |  |  | SGIT | cod | 1.08×10^-6^ | 1 |
| *SON* | 21 | 33543038 | SGIT | ncod | 1.57×10^-6^ | 1.57×10^-6^ |
| *ADARB1* | 21 | 45073853 | SGIT | ncod | 3.87×10^-7^ | 3.87×10^-7^ |
